# A method for analyzing the ERP associated with high frequency ANT DBS offset

**DOI:** 10.1101/2025.04.01.25324757

**Authors:** Herkko Mattila, Anna-Liisa Satomaa, Kai Lehtimäki, Jari Hyttinen, Sari-Leena Himanen, Jukka Peltola

## Abstract

**Objective:** We describe a sensor and source space electroencephalography (EEG) analysis method for clinically established high frequency (140 Hz) deep brain stimulation (DBS) of the anterior nucleus of thalamus (ANT). We demonstrate this by evaluating the EEG effects time locked to DBS in a single participant.

**Methods:** We used 1 s on and 5 s off stimulation on/off durations, providing a large number of trials. After artifact cleaning, the stimulation on-off EEG were averaged, and sensor and source space signals were determined and assessed.

**Results:** We found an event-related potential (ERP) time locked to the DBS stimulation offset. Source localization analysis revealed that the main sources of this ERP were localized in the limbic system (Papez circuit). Further, the sources in the anterior cingulate cortex were found to oscillate in synchrony with the ERP.

Conclusions

This study shows that time-locked EEG analysis related to monopolar bilateral 140 Hz ANT DBS stimulation is feasible.

**Significance:** The developed toolset reveals novel time-locked effects of high frequency ANT DBS on brain activity, which shows the future potential of sensor and source space analysis for DBS. The ERP findings might also prove useful in the optimization of epilepsy treatment with ANT DBS.

**Highlights:**

- We present a toolset for analyzing EEG effects during high frequency deep brain stimulation of the anterior nucleus of thalamus.
- Time locked to DBS offset, an event-related potential (ERP) occurred in EEG.
- Source analysis pointed to the limbic system and found synchronization of the anterior cingulate cortex with the ERP.

## 1. Introduction

Deep brain stimulation (DBS) targeted to the anterior nucleus of thalamus (ANT) is a treatment option for drug resistant epilepsy. The Stimulation of the Anterior Nucleus of Thalamus for Epilepsy (SANTE) trial has reported that ANT DBS reduced seizures by a median of 56% at two-year follow-up. At five-year follow-up, the median seizure frequency had further declined by 69% from baseline (Fisher et al., 2010; Salanova et al., 2015) and by 75% at seven-year follow-up (Salanova et al., 2021). The safety and efficacy of ANT DBS for the treatment of epilepsy has also been demonstrated in clinical practice (Peltola et al., 2023). Despite the group level efficiency of ANT DBS, certain patients benefit from it more than others. Knowledge of the factors predicting therapy outcome is, however, sparse. Therefore, finding a biomarker for therapeutic effectiveness would be very valuable.

The effects of DBS on brain function have been studied widely. The most frequently used methods are functional connectivity, EEG synchronization/desynchronization, changes in EEG power spectrum, evoked potentials, and functional magnetic resonance imaging (fMRI). Further, electroencephalography (EEG), stereo EEG (SEEG), and Local Field Potential (LFP) can also be applied (for a review see Fisher, 2023). However, due to a multitude of configurable DBS treatment parameters, the comparison of different studies is not a straightforward task. For example, the stimulation target, the selected electrodes (bipolar or monopolar), and the stimulation current and frequency can all vary, and either continuous or intermittent stimulation can be used. In addition, the measurements may be done in sleep or wake, intra- or extracranially, intra- or postoperatively, or for longer periods of time utilizing the chronic sensing properties of most modern DBS devices. Moreover, the study paradigms have varied from animal models to experimental and clinical research. Importantly, knowledge of the EEG effects on the clinically established high frequency ANT DBS is limited. Indeed, there have been only a few attempts to study brain responses to DBS at the source level (Zumsteg et al., 2006a). From a clinical perspective, however, an understanding of DBS effects on brain function and finding the location of the EEG sources are important, as they might help to better understand the therapeutic efficiency of DBS.

ANT DBS is known to induce evoked cerebral responses. To our knowledge, previous EEG studies that have addressed the evoked cerebral responses have used a 2 Hz DBS frequency (Zumsteg et al., 2006b, 2006c, 2006a). In a recent fMRI study, it was reported that high frequency (145 Hz) vs low frequency (30 Hz) ANT DBS produces quite different cortical responses (Middlebrooks et al., 2021). Therefore, it is unclear whether 140 Hz stimulation, which is typical for the therapeutic use of ANT DBS, generates cortical evoked responses. Furthermore, in previous studies on evoked responses there have been other methodological shortcomings. For example, in the study by Zumsteg and co-workers (Zumsteg et al., 2006a), the number of EEG electrodes (27 channels) was low, the EEG electrode positions were not digitized, and individual MRI data were not used.

In this methodological proof-of-concept study, we present a new approach for assessing EEG responses to ANT DBS stimulation at sensor and source levels. To further demonstrate our methods, we investigate whether high frequency (140 Hz) stimulation induces time-locked cerebral responses in EEG, which we denote ERPs. Finally, we discuss the potential of the method in the optimization of epilepsy treatment with ANT DBS.

## 2. Materials and methods

### 2.1. Study participant, DBS implantation, and therapeutic DBS procedure

In this methodological work, we present a case study of a pilot patient. The study was conducted in accordance with the Code of Ethics of the World Medical Association (Declaration of Helsinki) and the Ethical Committee of Pirkanmaa Hospital District approved the study (R17132). Written informed consent was obtained from the participating patient.

DBS electrodes with stimulator targeted to the ANT were implanted to a young adult patient with drug resistant epilepsy. The patient had the seizure-onset-zone in the right temporo-occipital region, with cortical dysplasia as the etiology. The possibility of surgical resections was ruled out based on the findings of preoperative investigations. On average, the patient had more than 10 monthly focal impaired awareness seizures (FIAS) despite the patient being prescribed three anti-seizure medications (ASM). As the patient preferred ANT DBS over vagal nerve stimulation, ANT DBS stimulation was chosen. Medtronic 3389 leads (Figure S.1) and a Medtronic Activa PC implantable pulse generator (IPG) were implanted under general anesthesia using the Leksell stereotactic system. The target was preoperatively defined in 3T MRI, enabling direct visualization of the borders of the ANT and the mammillothalamic tract (Möttönen et al., 2015). The surgical technique has been previously described in detail (Cukiert and Lehtimäki, 2017; Lehtimäki et al., 2016).

A postoperative helical computed tomography (CT) scan (Revolution GSI, GE Medical Systems, Boston, MA, USA) was performed to define the actual positions of the implanted DBS electrodes. The postoperative CT scan was then co-registered with the preplan magnetic resonance (MR) images. Finally, SureTune 3.0 software (Medtronic, Minneapolis, Minnesota, USA) was used to create a 3D image visualizing the positions of the stimulation electrodes relative to the segmented ANT (Järvenpää et al., 2020). The same software was also used to simulate the monopolar electric field that the stimulus parameters used in this study would induce in homogenous tissue (Figure 1).

**Figure 1.**
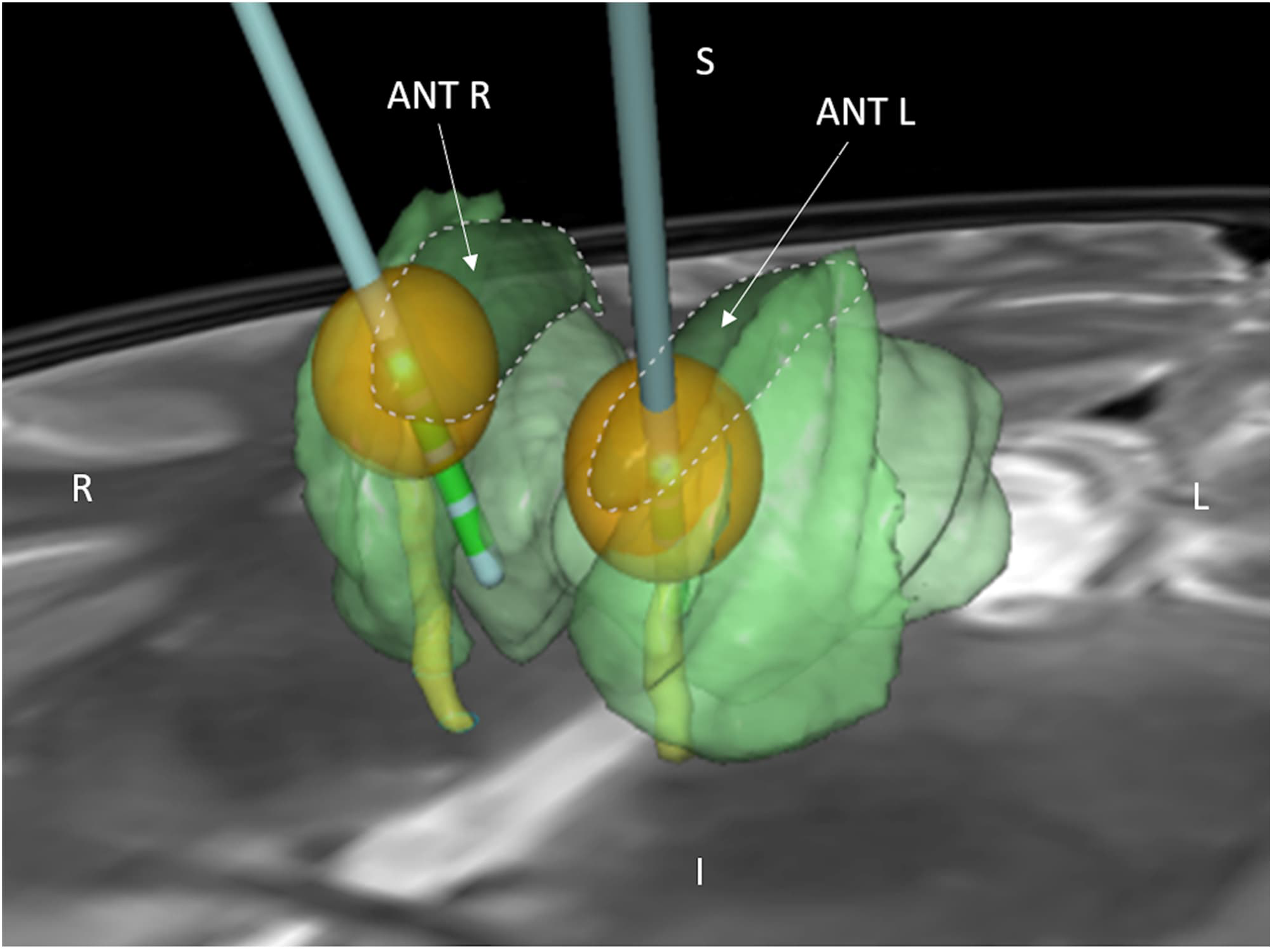
The stimulation electrodes (light blue with green contacts) implanted into the left and right ANT (ANT L and ANT R, respectively). The yellow fibers represent the mammillothalamic tract.

The following stimulation protocol for the patient’s epilepsy treatment was used: pulse width 90 µs, voltage 5 V on the left and right sides, with a stimulation frequency of 140 Hz. The thalamus was stimulated bilaterally with a bipolar configuration. Electrode contacts 2 and 3 were cathodes and contact 0 was anode on the left side of the brain. Electrode contacts 10 and 11 were cathodes and contact 8 was anode on the right side of the brain. The stimulator activity was 1 minute on and 5 minutes off periodically. These parameters were chosen due to insufficient seizure response with a primary SANTE type stimulation protocol (Fisher et al., 2010). The patient was classified as a responder (>50% seizure reduction) using these bipolar parameters.

Although the therapeutic stimulation protocol of the patient was bipolar, a monopolar stimulation protocol was applied during the study recordings. The orange spheres represent the simulated electrical fields of the monopolar 5 V stimulation through contacts (cathodes) 3 and 11, respectively; the IPG was anode. Orientation marks in the image: Superior S, Inferior I, Left L and Right R.

### 2.2. EEG recording setup and visual analysis

When the patient first arrived at our laboratory for the DBS test session and the concurrent EEG recording, the DBS stimulator was turned off. EEG was recorded with 64-channel active electrode cap (actiCAP slim/actiCAP snap, Brain products GmbH, Gilching, Germany). The international 10-10 positioning of the EEG electrodes was used, and the electrode impedances were fixed below 5 kΩ. A reference electrode was placed at FCz and a ground electrode at Fpz. Two bipolar electrooculogram (EOG) channels were recorded; one to record horizontal eye movements and the other to record vertical eye movements (Häkkinen et al., 1993). A two-lead ECG was recorded with one electrode in the right shoulder and one in the left lower rib. In addition, a sleep mattress sensor (EMFIT Ltd, Vaajakoski, Finland) was positioned under the patient’s mattress to record breathing, body movements, and ballistocardiogram. All the electrodes were connected to a Neurone Tesla amplifier (Bittium Biosignals, Oulu, Finland). The sampling rate was 20 kHz, and the amplifier frequency range was from 0 Hz (DC) to 3500 Hz. The dynamic range of the channels was ±430 mV. The high sampling rate and wide dynamic range were selected to make it easier to handle the DBS artifact in the EEG recordings (Lio et al., 2018).

The patient had smoked a cigarette 2.5 hours before and consumed two cups of coffee 4 hours before the EEG recording. In addition, the patient had had an epileptic seizure two hours before the test session began. The seizure was typical FIAS, with a short period of unawareness and full recovery after 5 minutes. The recording session took place in a quiet sleep laboratory room with dim lightning. During the recording, the patient was seated in a semi-sitting position on a hospital bed. The position was made as comfortable as possible to avoid any tension in the neck and shoulders. Next, the patient was given the following instructions: “Close your eyes and relax. You may daydream, but do not think about any specific problem or issue. Do not sleep.” In addition, the patient was asked to notify the researchers if he felt a seizure coming on during the EEG recording. When the stimulator had been off for approximately one hour, and while the stimulator continued to be completely off, a 5-minute baseline EEG sample was recorded.

After the baseline EEG sample was recorded, several different DBS test protocols with variable stimulation parameters were carried out. However, in the present study only one of the test protocols, the bilateral 140 Hz monopolar stimulation, was evaluated. More specifically, the stimulation setup for the purposes of the present study was as follows: the stimulating electrode contacts of the DBS electrodes were 3 and 11 (Figures 1 and S.1). The contacts were inside the ANT. The stimulation was constant voltage and monopolar so that the IPG was anode and the electrodes 3 and 11 were cathodes. The impedances of electrodes 3 and 11 were 1135 Ω and 1284 Ω, resulting in stimulation currents of approximately 4.4 mA and 3.9 mA, respectively, through the electrodes. The pulse width was 90 us, voltage 5.0/5.0 V, and frequency 140 Hz. Regarding the technical specifications of the Activa PC IPG, the left and right hemispheres received stimulation alternately, not simultaneously. This means that although the left and right ANT were both stimulated at a frequency of 140 Hz, there was approximately a 3.57 ms latency between the left- and right-sided stimulation pulses, resulting in a total brain stimulation frequency of 280 Hz. The stimulator was on for 1 second and off for 5 seconds periodically for a total of 10 minutes. Consequently, the EEG recording used in the present study comprised 100 “stimulator on” and 100 “stimulator off” segments.

BrainStorm software, version 20-may-2019 (Tadel et al., 2011), was used in the visual analysis, signal processing, source modeling, and presentation of the EEG results. The software is documented and freely available for download under the GNU General Public License (http://neuroimage.usc.edu/brainstorm).

The EEG signal was visually screened using display filtering from 0.5 Hz to 100 Hz. Three EEG segments with electromyogram (EMG artifacts) exceeding 100 μV were excluded from further analysis. In addition, an experienced clinical neurophysiologist (AS) ensured that the patient had stayed awake during the sessions and estimated the amount of interictal abnormal activity during the baseline and test sessions to be equal.

### 2.3. EEG electrode location digitization

After the EEG recordings, the positions of the EEG electrodes as well as the patient’s head points were digitized to the patient’s individual MR images. The digitization was done with stereotactic infrared camera (Polaris Vicra, NDI, Ontario, Canada), using the digitization tools of a navigated transcranial magnetic stimulation (nTMS) device (Nexstim NBS System 4, Nexstim, Helsinki, Finland). The positions of three fiducial points (nasion, left and right ear crus of helix), the EEG electrodes, and more than 100 digitized head scalp points were digitized and stored for later use.

### 2.4. Signal preprocessing

EEG was 0.5 Hz to 100 Hz bandpass FIR filtered (Brainstorm software filter version 2019), with transition band 0 Hz, using a 60 dB Kaiser filter to remove the 140 Hz stimulation artifact, high frequency noise, and DC offset.

For artifact cleaning, 64 independent component analysis (ICA) components were calculated using the InfoMax algorithm (Bell and Sejnowski, 1995; Makeig et al., 1996). The EEG bandpass frequency range was used, i.e., the calculation was made on a continuous file; and the input signals were resampled to 2000 Hz. The ICA components to be removed were selected by visual evaluation of the time series and topography maps of the ICA components and by inspection of the resulting EEG distortion of the selected ICA components. The ICA was then calculated independently for the baseline run and for the DBS test protocol run. The artifact cleaning is described in more detail in chapter 2.4.1 and in the supplementary file.

The last stimulus pulse in all the 1 second stimulation periods was marked as a stimulation off event to be used later in the data epoching, which is explained in more detail in chapter 2.4.2. and in the supplementary file. Finally, the sampling rate of the EEG signal was downsampled from 20 kHz to 2 kHz.

#### 2.4.1. EOG, EMG, and stimulation artifact cleaning

The ICA was used to clean EOG and small amplitude EMG artifacts of the individual EEG channels. The first and the last pulse in the stimulus train induced their respective DBS stimulus filtering artifacts, which were removed by nine ICA components. The effects of these components are presented in Figure S.2 in the supplementary file. The topography and a sample of the time series of the ICA components are presented in Figure S.3.

#### 2.4.2. Epoching and averaging

As the stimulus onset and offset caused filter ringing (Figure S.2), the epochs were fixed to begin −500 ms and end +1000 ms relative to the stimulation off event markers in the further analysis. As a result, 100 epochs each of 1500 ms in duration were created.

Despite the artifact cleaning, some residual artifacts were found and five out of 100 epochs were rejected from the final average, resulting in 95 epochs. The 95 artifact-free epochs were averaged together to extract the event-related response. Moreover, it was confirmed that this ERP-like waveform was not generated by the stimulation or the filtering of the artifacts (Figures S.4 and S.5).

#### 2.4.3. Remainder sinc-shaped artifact in the averaged signal

Some filtering-related sinc-shaped artifacts remained (Figure S.6) and, despite several attempts, this artifact could not be removed. Therefore, the time range from 0 ms to 50 ms was excluded from the final analysis.

### 2.5. Source localization

#### 2.5.1. MRI segmentation

The surgical preplan MR images were imported to FreeSurfer 6.0.0 software (http://surfer.nmr.mgh.harvard.edu/), which was used for cortical reconstruction and volumetric segmentation (Dale et al., 1999; Dale and Sereno, 1993; Desikan et al., 2006; Fischl et al., 2004b, 2004a, 2002, 2001, 1999b, 1999a; Fischl and Dale, 1999). FreeSurfer then reconstructed the surfaces using default parameters. As a result, triangulated surface models of the cortex, white matter, and subcortical structures were obtained. The results were visually confirmed. The software also created cortical and subcortical parcellations, which can be used as atlases.

#### 2.5.2. Forward modeling

Next, the original preplan MR images and the triangulated surface models generated by FreeSurfer were imported to Brainstorm software. The previously digitized EEG electrode positions were then co-registered to the patient’s MR images in Brainstorm using the fiducial points as references. The digitized scalp points were used to refine the co-registration. Finally, to compensate EEG electrode height (5 mm), the positions of the EEG electrodes were projected to the scalp. Moreover, using OpenMEEG software (Gramfort et al., 2010; Kybic et al., 2005), which is integrated in Brainstorm, a three-layer boundary element method (BEM) head model was created. Default software parameters, including relative conductivities (for the scalp, skull, and brain 1.0000, 0.0125, and 1.0000, respectively) were used.

#### 2.5.3. Source modeling

Brainstorm software was used to create a distributed source model of the averaged EEG epochs, which entails the software inserting a dipole in the vertex of every triangle of the triangle tessellated cortex surface. The original surface model of the cortex comprised 267 376 vertices, but for source reconstruction purposes the number of cortical vertices was resampled to 15 000 in the Brainstorm software. The dipoles were oriented normal to the cortical surface. By default, the Freesurfer-reconstructed cortex was a closed surface, as the surface inverse solution method requires a closed surface. Therefore, a part of the tessellated surface comprised deep medial areas of each hemisphere in addition to the cerebral cortex, which prompted us to test both the cortical surface model and the volume model. The result was that some activity was found in deep subcortical regions in addition to the cortex, which advocated the creation of a mixed source model instead of a cortical source model (Attal et al., 2009; Attal and Schwartz, 2013). Briefly, this means that in addition to the cortical surface, the software was allowed to distribute the dipoles to certain subcortical nuclei, as presented in the supplementary file (Figure S.7). The activity of these deep subcortical sources is not reported in this study, as the level of scientific evidence based only on a single patient and one stimulation condition would not have been sufficient. For the methodological evaluation of the source localization, however, the statistically significant activity in the deep brain areas is presented in the supplementary file (Figure S.8).

#### 2.5.4. Source computation

The sources were computed for the timeline of the averaged EEG epochs in the BrainStorm software using the dynamic Statistical Parametric Mapping (dSPM) method (Dale et al., 2000). The baseline EEG file was used to compute the noise covariance matrix. The covariance matrix was used for the noise de-whitening during the source reconstruction process. Default parameters were used (compute sources 2018: depth weighting: order 0.5, maximal amount 10; regularize noise covariance: 0.1; regularization parameter 1/λ: signal-to-noise ratio 3). Finally, the dSPM source maps were scaled by multiplying with the square root of the number of the averaged epochs to maintain the variance of one under the null hypothesis. It should be noted that all the EEG data of the baseline recording were assumed to represent the noise signals in the dSPM calculations. This means that Z-score in the dSPM activity maps represents deviation from this baseline activity. The Brainstorm software workflow is explained in more detail on the software tutorial pages https://neuroimage.usc.edu/brainstorm/Tutorials/Workflows), (https://neuroimage.usc.edu/brainstorm/Tutorials).

### 2.6. Statistics

The normal distribution of the ERP amplitude variability in sensor space was studied using the Shapiro-Wilk statistical test. For the testing, the amplitude values of the Oz EEG channel were extracted from trials at the time points of N80 and P120.

For the source estimation, the dSPM method was used to estimate the statistical significance of the source activity. The dSPM compares the activity of each dipole to the baseline activity of that specific dipole. The SPM method uses random field theory (RTF) to handle the multiple-comparison problem.

### 2.7. Availability of research data

Research data is conditionally available from the authors with a formal data sharing agreement.

## 3. Results

### 3.1. Stimulation offset Event-Related Potential

The result of the averaged epochs is presented in Figure 2. The obtained cortical response resembles an ERP, which has positive (P) and negative (N) peaks in the time range of 50 ms to 275 ms. The time point 0 ms is the peak of the last stimulus pulse. The following peaks can be discerned: N50, N80, P120, P165, N190, N240, and P275. The peaks N80 and P120 present the highest amplitudes. The ERP distribution in the sensor space is shown in Figure 3. Using visual assessment, the ERP response appears symmetric between the left and right hemispheres, and its amplitudes seem higher in the posterior than in the frontal areas.

**Figure 2.**
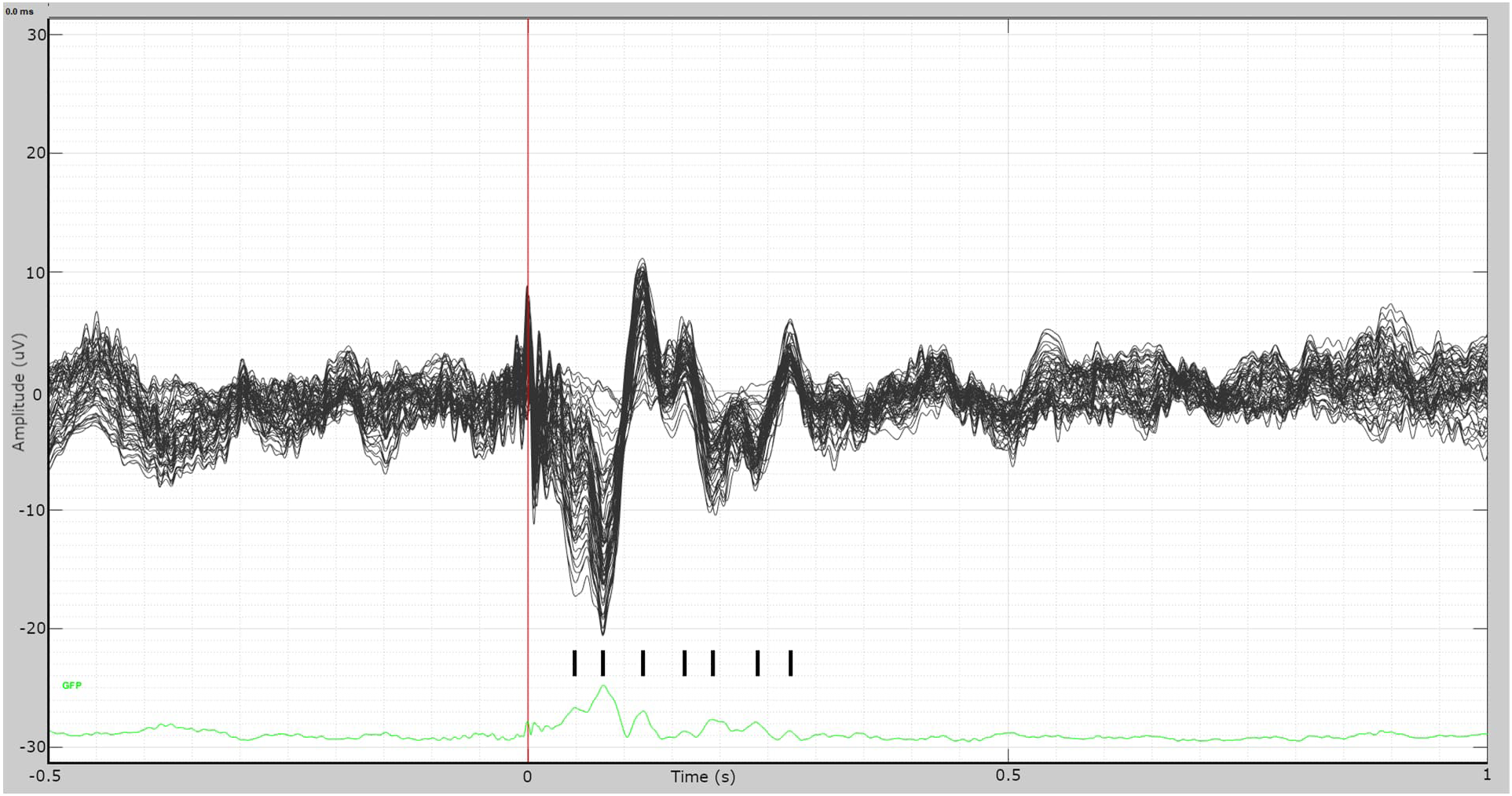
Butterfly plot of the 64 EEG channels. Averaged epochs in the time range from −500 ms to 1000 ms are presented. The vertical scale is ±30 µV. The ERP-like waveform can be seen in the time range from 50 ms to 285 ms. The vertical red bar shows the time point of 0 s, which is the time point of the last stimulus pulse. The green curve at the bottom presents the global field power (GFP) (Lehmann and Skrandies, 1980; Skrandies, 1990). The black tick marks above the green GFP represent the peaks of the ERP. The peaks are approximately N50, N80, P120, P165, N190, N240, and P275, where N denotes negative (downward) and P positive (upward) peaks, and number denotes the latency in milliseconds.

**Figure 3.**
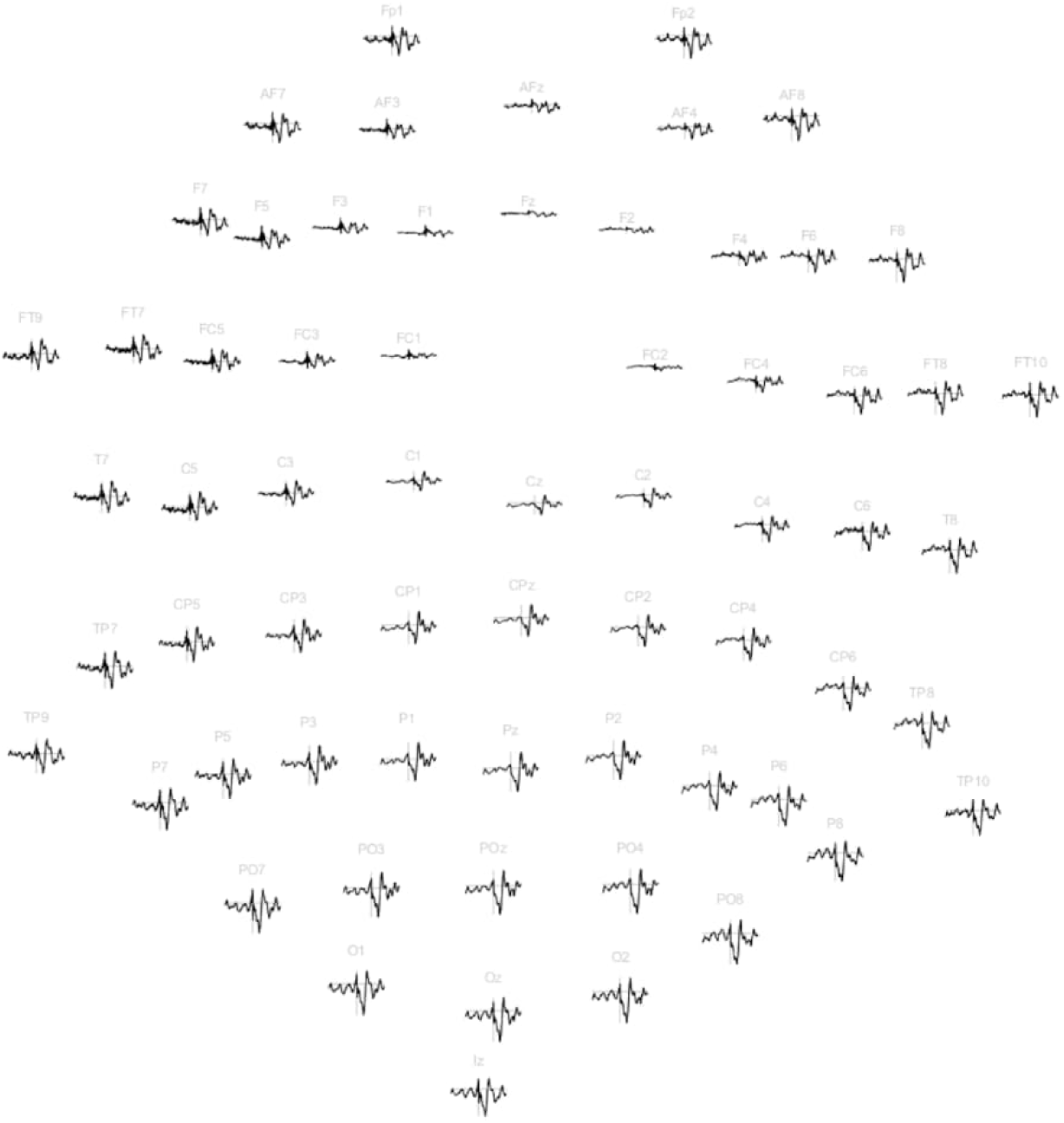
The 2D-layout of the ERP response in the time range of −320 ms to +320 ms. The maximum amplitude is 21 μV. Reference electrode FCz.

In Figures 4 and 5, the amplitude variances of N80 and P120 are presented in the Oz EEG channel. The variability of the amplitude is normally distributed, but no clear time trend can be seen.

**Figure 4.**
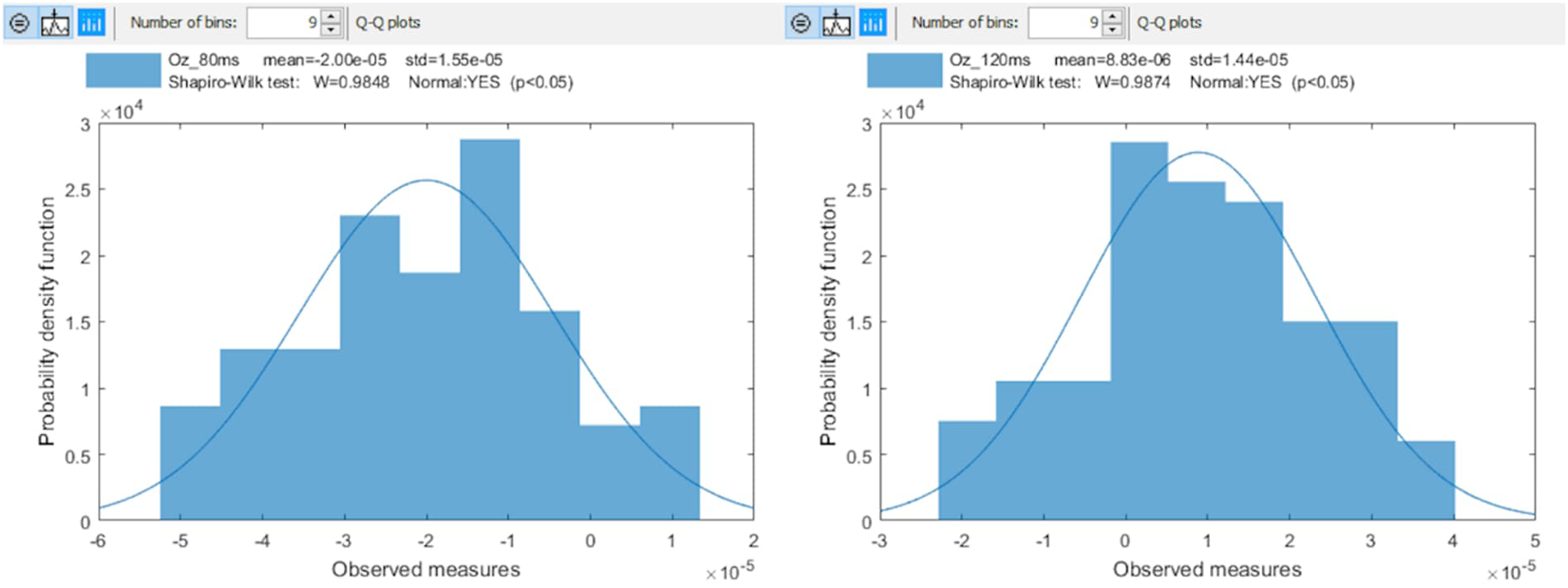
Histogram plots of the N80 and P120. ERP components measured from the EEG channel Oz in the epochs from 1 to 95. The x-axis presents the amplitude as 10^−5^ volts. Statistically, the variabilities of N80 and P120 amplitudes are normally distributed.

**Figure 5.**
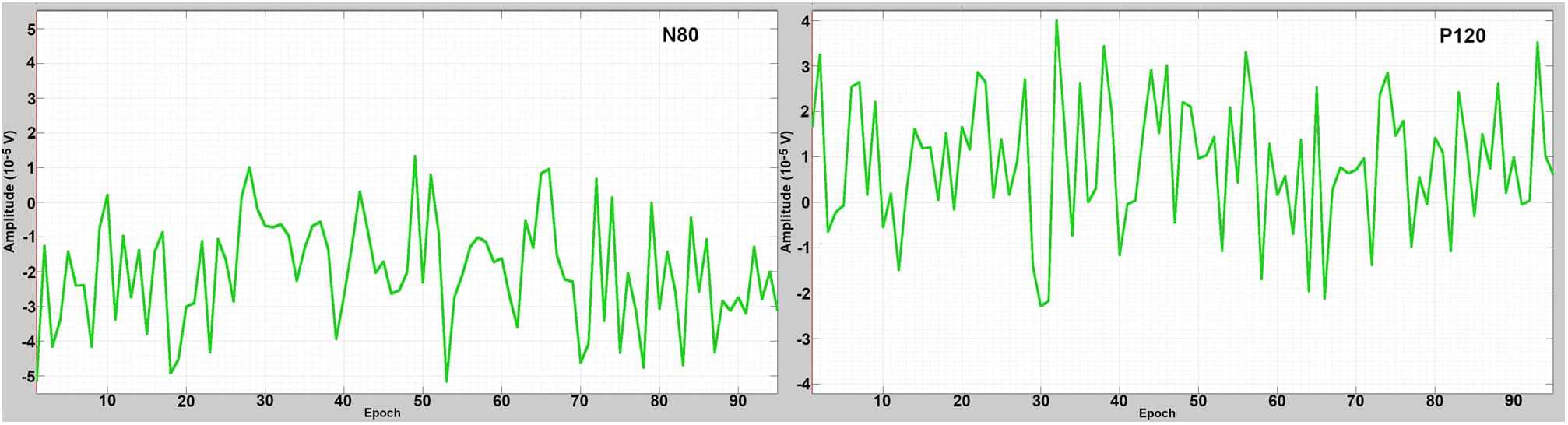
Time variability of N80 (left image) and P120 (right image) amplitude in Oz. The horizontal axis presents the epochs from 1 to 95 and the vertical axis is the amplitude range, which is ±58 µV in the left image and ±47 µV in the right image. No time trend in amplitude variation of the N80 or the P120 was observed.

### 3.2. Cortical sources of the ERP

In Figures 6 and 7, which represent the cortical sources, the cortical surface gyrification is maximally smoothed (100%) for the visualization of sulci. The lighter and darker shades of gray represent the cortex and the sulci, respectively. In figures 6 and 7, the colormap representing the activity is based on a statistical threshold. In addition, only the source clusters of 20 dipoles or more are presented.

**Figure 6.**
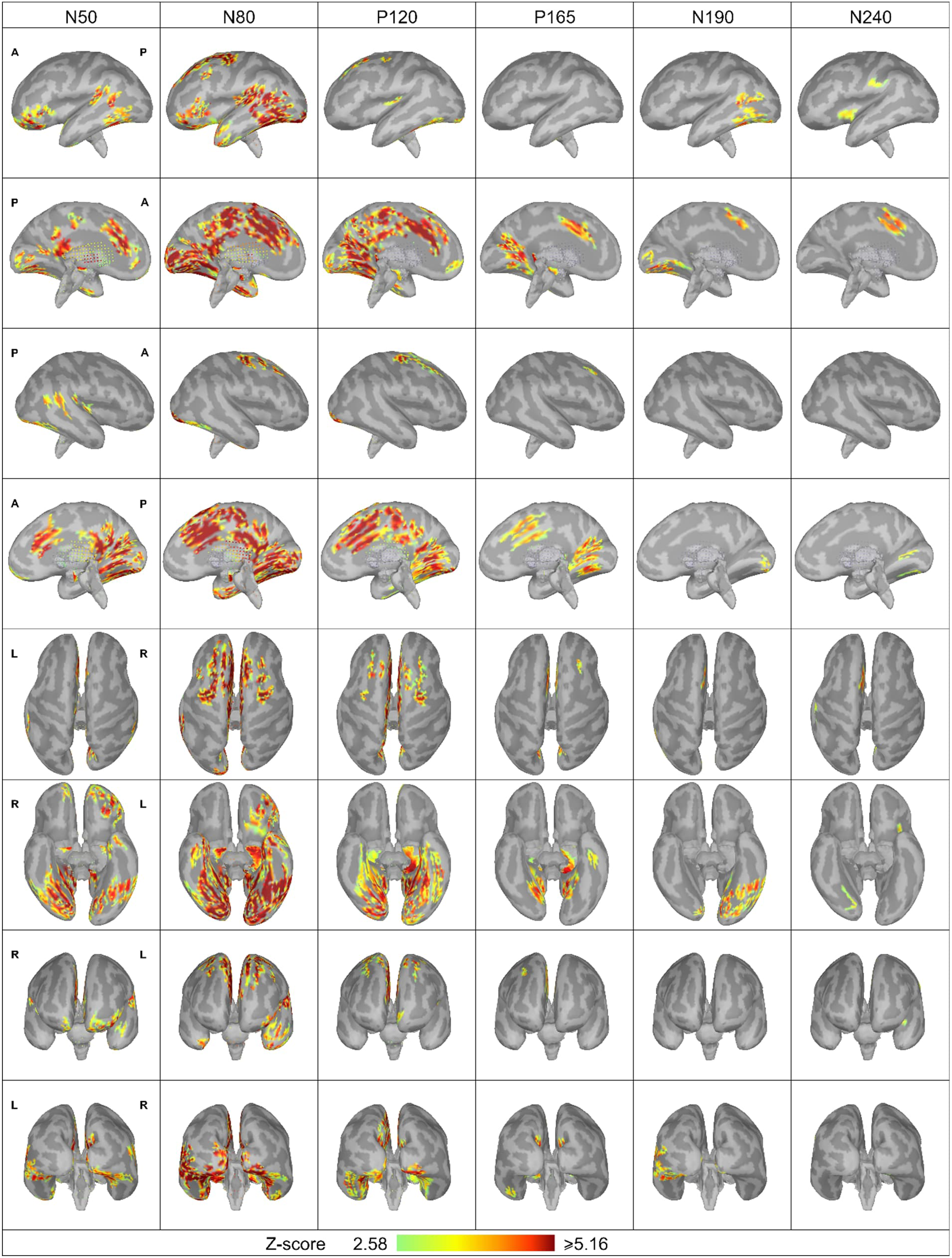
Source maps of the cortical ERP components. The cortical surface is smoothed 100%. Here, lighter gray represents the gyri, and darker gray represents the sulci. Only activity with p<0.01 is presented (Z-value > 2.56). First row: left hemisphere lateral view, second row: left hemisphere medial view, third row: right hemisphere lateral view, fourth row: right hemisphere medial view, fifth row: view from the top of the brain, sixth row: view from the bottom of the brain, seventh row: anterior-posterior view, eighth row: posterior-anterior view.

**Figure 7.**
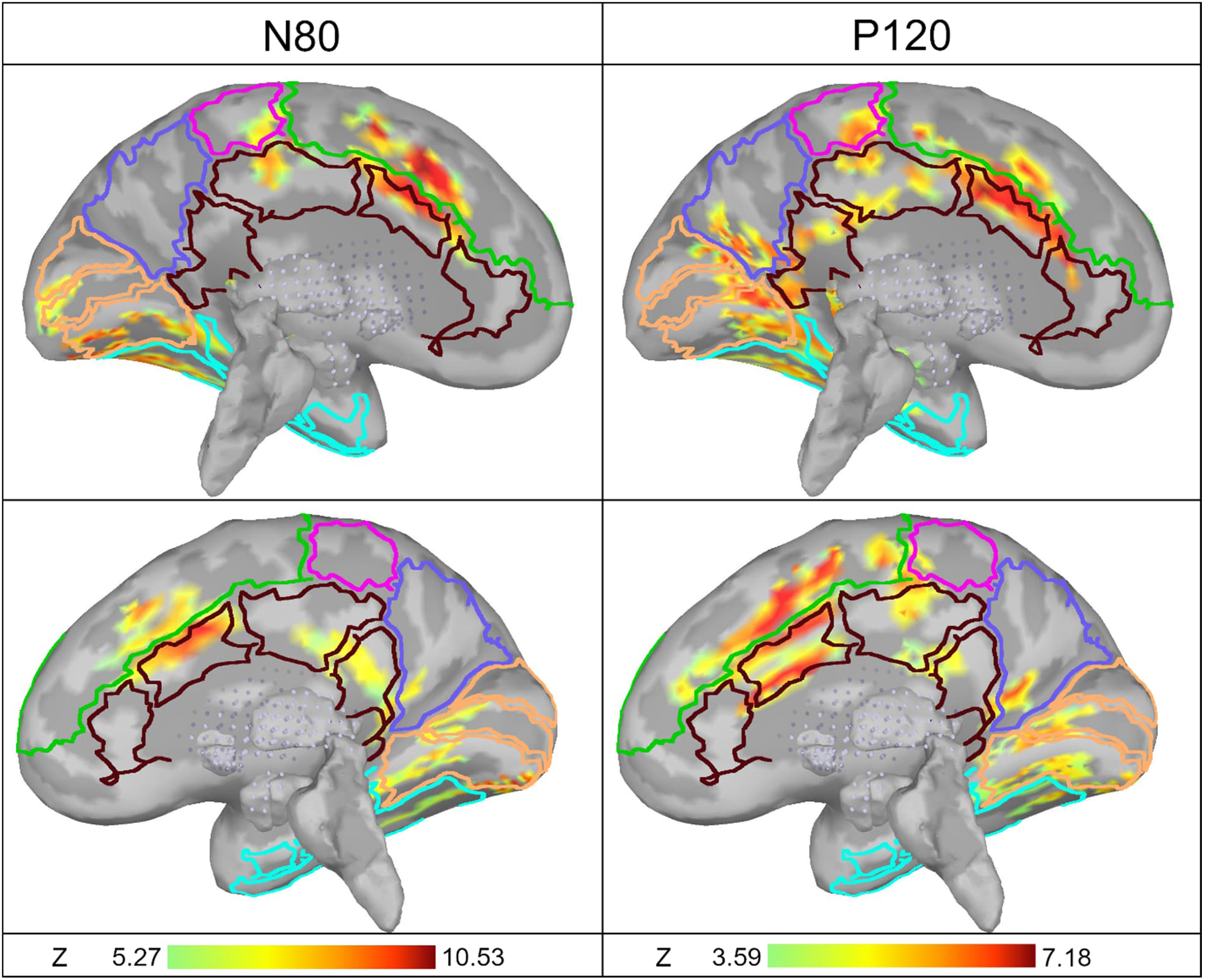
The maximal activation of the medial surfaces of the left and right hemisphere during N80 (left images) and P120 (right images) components. The color scale threshold is 50% of maximum value and the cortical surface is smoothed 100%. Lighter gray represents the gyri and darker gray the sulci. The minimum size of the activated area is limited to 20 dipoles. The color scale is from 5.27 to 10.53 (Z-distribution) in N80 and from 3.59 to 7.18 in P120. Only activity with p<0.001 (Z>3.3) is presented. The borders of the Desikan-Killiany cortical atlas are also projected over the cortex. The regions are color coded: green, medial frontal region; brown, cingulate region (rostral anterior cingulate, caudal anterior cingulate, posterior cingulate, and isthmus cingulate); magenta, central region (paracentral); blue, parietal region (precuneus); orange, occipital region (lingual, pericalcarine and cuneus); turquoise, temporal region (entorhinal, parahippocampal and fusiform).

Figure 6 visualizes the main findings. First, the activation of the homologous cortical areas of the two hemispheres were not synchronous. Second, even though the activations were asynchronous, the cortical areas activated were almost the same in each hemisphere. The activity sources were found in the following areas: the frontal cortex (superior frontal, rostral middle frontal, caudal middle frontal, pars triangularis), the orbitofrontal cortex (frontal pole, lateral orbitofrontal, pars orbitalis), the temporal cortex (temporal pole, insula, superior temporal, banks of the superior temporal sulcus, middle temporal, inferortemporal, fusiform, parahippocampal, entorhinal), the central cortex (precentral, paracentral), the parietal cortex (supramarginal, inferior parietal, precuneus), the occipital cortex (lateral occipital, lingual, pericalcarine, cuneus), and the cingulate cortex (rostral anterior cingulate, caudal anterior cingulate, posterior cingulate, isthmus cingulate). In other words, the sources of the ERP were found in the frontal, temporal, parietal, and occipital regions as well as in the cingulate cortex in the medial surface of both hemispheres. Pars orbitalis and pars triangularis were found to be active only in the left hemisphere. The anterior and superior part of the temporal lobe as well as the most posterior temporal lobe adjacent to the parietal lobe were more active in the left than in the right hemisphere.

The maximal activations of the medial surface during the highest peaks, N80 and P120, are presented in detail in Figure 7. The color scale is scaled to the local maximum of N80 (Z-value 10.53) and P120 (Z-value 7.18). The strongest activity was found in the caudal anterior cingulate and the medial frontal region.

### 3.3. Time series of the anterior cingulate sources

Figure 8 presents the mean time amplitude signals of the dipole sources in the regions of the left and right caudal anterior cingulate cortex. The activity oscillates and the activity maximums coincide with the ERP peaks N50, N80, P120, P165, N190, N240, and P275.

**Figure 8.**
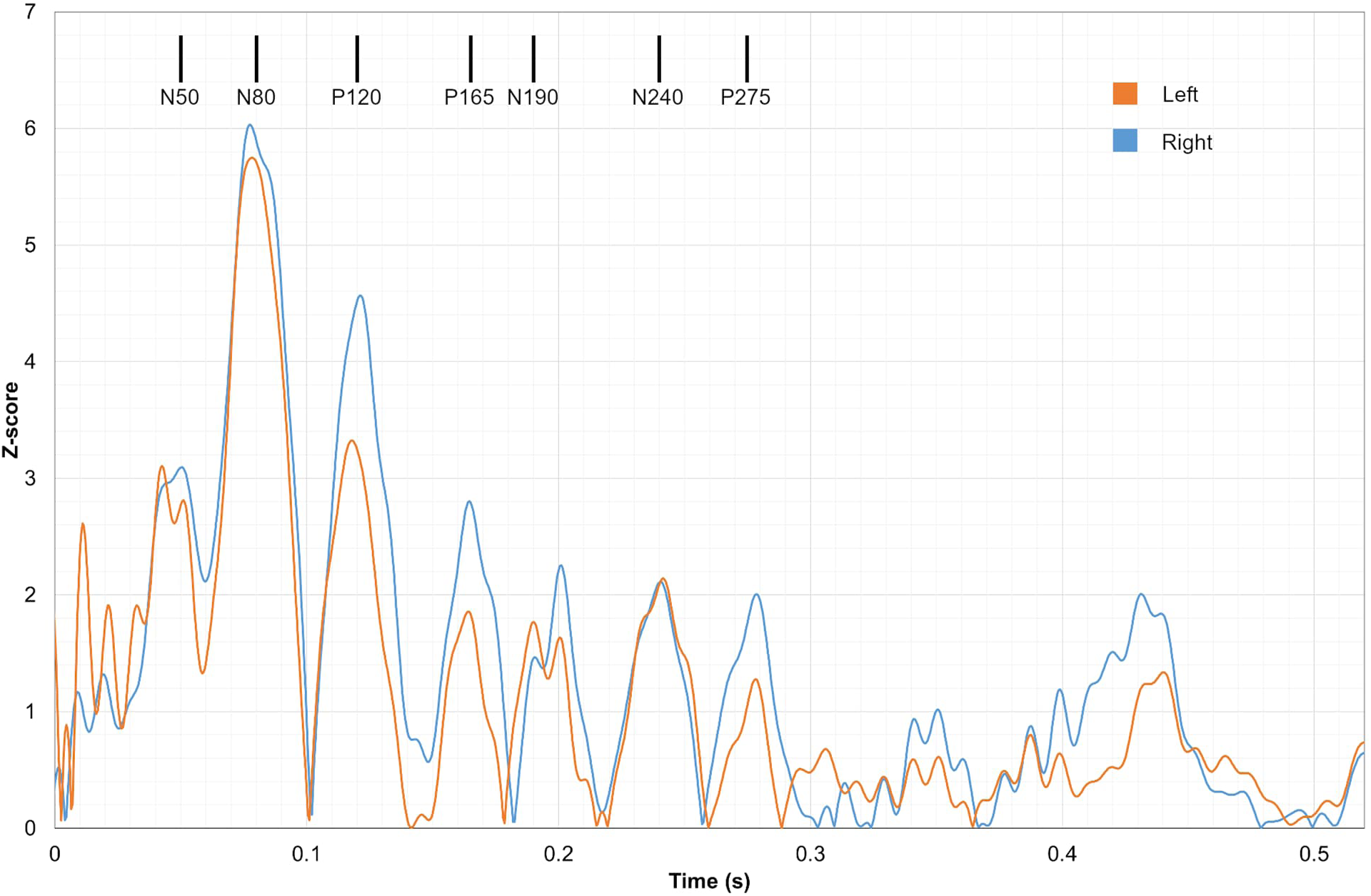
The mean time-amplitude signals of the dipoles in the left and right sides of the caudal anterior cingulate cortex. Z-score of the amplitude is presented. Black vertical bars denote the time points of the ERP components N50, N80, P120, P165, N190, N240, and P275. The caudal anterior cingulate cortex oscillates in synchrony with the ERP.

## 4. Discussion

In this study, we present a novel method for analyzing the EEG responses related to monopolar, bilateral, high frequency (140 Hz) ANT DBS in source and sensor space. The method includes stimulus design, denoising and stimulus artifact removal, DBS ERP epoch formation, and inverse source localization and source level signal determination. The special stimulus on-off timing and sufficient stimulation artifact removal enabled the analysis of the EEG.

Demonstrating the feasibility of the presented method, a new ERP-like cortical response time locked to ANT DBS offset was found. The resulting ERP was studied in both sensor space and source space using the inverse EEG imaging method. Until now, the methods applied in DBS EEG source localization studies have not used the same DBS stimulation parameters as commonly used to treat epilepsy. This is of importance, as the acute brain activation patterns of the ANT DBS have been found to depend on the frequency of the stimulation in fMRI studies (Middlebrooks et al., 2021). Indeed, to our best knowledge, our DBS EEG study is the first study to evaluate brain activity and the dynamics of the stimulus response time-locked to DBS using the same stimulation parameters (high frequency, bilateral monopolar stimulation) commonly applied in the ANT DBS treatment of epilepsy.

The observed symmetric distribution of the ERP over the scalp in sensor space implies that the main sources of the ERP are probably somewhere in the midline of the brain. Source localization results support this finding. Although source activity was distributed over different regions of the brain (frontal, temporal, parietal, and occipital), the strongest source activity was found on the medial surface of the brain, that is, the cingulate gyrus and inferior areas of the superior frontal gyrus.

Despite strong bilateral synchrony for most regions, some asymmetry between the left and right hemispheres was also found. For example, in the insula and entorhinal cortex, the source activity was found only on the left side. Moreover, time series analysis of the sources revealed that the caudal anterior cingulate gyrus activity oscillated in synchrony with the ERP.

Our findings in sensor and source space are broadly in agreement with previous EEG studies conducted by Zumsteg et al. (Zumsteg et al., 2006b, 2006c, 2006a). They found that ANT stimulation resulted in activation of the ipsilateral anterior cingulate gyrus, insula, parietal operculum, inferior temporal cortex, and mesial temporal cortical areas. Furthermore, in line with the findings of the present study, Zumsteg et al. also found the caudal anterior cingulate cortex oscillated in synchrony with ERP (Zumsteg, Lozano, Gregor, et al., 2006). Importantly, they showed the intra-subject reproducibility of DBS-related ERP in a low frequency stimulation setup. In contrast to previous studies which applied only 27 channels, we had 64 EEG channels, improving source space accuracy greatly.

One of the main anatomical connections of ANT is known to be with the anterior cingulate cortex (Child and Benarroch, 2013), which is a part of the medial limbic circuit (Papez circuit). Further, the DBS-related activity of limbic networks has been demonstrated in EEG and fMRI studies (Gibson et al., 2016; Middlebrooks et al., 2021, 2020, 2018; Zumsteg et al., 2006a). Therefore, it was not unexpected that in our study ERP source activity was also found to be high in these regions.

Unfortunately, ANT DBS does not benefit all patients. It is not known, however, whether good therapeutic response is associated with the individual characteristics of the patient or the type of epilepsy, or whether it relates to the stimulation setup applied. Regarding the intrinsic features of the patient, there is little knowledge of which factors might associate with treatment outcomes. For example, poor executive (Järvenpää et al., 2018) or cognitive (Kaufmann et al., 2020) function and seizures within the Papez circuit (Middlebrooks et al., 2018) have all been linked to poor or favorable therapy responses. From the perspective of brain function, it has been suggested that intact connectivity between the ANT and Default Mode Network (DMN) (Raichle et al., 2001) induced by ANT stimulation is essential for DBS effectiveness (Middlebrooks et al., 2018).

Recently, Aiello et al. (Aiello et al., 2023) found that ANT DBS responders have stronger EEG theta oscillations, higher theta power in the ANT, and stronger ANT to scalp connectivity than non-responders. The exact stimulation site is known to be important (Guo et al., 2021; Möttönen et al., 2015). In addition, while modern DBS stimulators offer several options to adjust the stimulation settings, enabling highly customized stimulation setups, there are no guidelines on how to optimize the stimulator settings. Therefore, in many cases the optimization of epilepsy treatment with ANT DBS requires patient level adjustment of the stimulus parameters. This is usually done through trial and error, and the efficacy of the treatment may be evaluated after weeks or even months.

Biomarkers based on various neuroimaging modalities, such as EEG, might help to accelerate this process by prognosticating the clinical response or by helping to optimize the stimulator settings individually.

In comparison to other neuroimaging modalities, EEG is an inexpensive, non-invasive, and readily available clinical method for everyday use. There is, however, no gold standard method to measure the time-locked EEG effects of ANT DBS. Previous studies that have evaluated the EEG effects of ANT DBS have applied relatively simplified test protocols and, importantly, low frequency stimulation (Zumsteg et al., 2006b, 2006a). As it is known that different DBS stimulation frequencies have different effects on brain responses (Middlebrooks et al., 2021), it is important that clinically established DBS setups (high frequency) are studied. The method we describe here was applied on high frequency ANT DBS and revealed an ERP. This ERP and the assessment of EEG responses at source and sensor levels may provide novel tools in the search for biomarkers. Regarding the clinical aspect, further studies are needed.

### 4.1. Limitations of the study

DBS artifact and its removal are major issues in DBS EEG measurements (Lio et al., 2018). We cleaned the stimulation artifacts by using a low-pass filter and by removing certain clearly artifact-related ICA components from the data. This led to sufficient, but incomplete DBS artifact removal. The ICA was also used to remove EMG- and EOG-related artifacts. The corresponding author selected all the removed ICA components by visual inspection and according to rather subjective decision making. Some of the decisions required compromising between artifact removal and deformation of the EEG. In this study, the noise covariance matrix was computed from the baseline data. We conducted the ICA cleaning separately at the baseline and the at the stimulation on/off run. However, because the ICA does not necessarily remove all the artifacts completely, the residual artifacts may have an effect on the estimated noise levels, and thereby on source localization.

Moreover, in the present study, only 64 channels of EEG were available, whereas a larger number of channels would have provided a smaller localization error in source space (Akalin Acar and Makeig, 2013). However, we demonstrated that source space imaging with 64 channels was accurate enough to conform to the details of the Desikan-Killiany atlas.

Selected subcortical structures were also included in the source model, but the results (supplementary file, chapter 2) were not reported, as the reliability of the scalp EEG in detecting the activity in deep subcortical structures is a matter of debate (Fahimi Hnazaee et al., 2020).

In the interpretation of the ERP results, a major weakness is that only one patient was studied and only one run in ERP measurement mode was performed. Therefore, the study lacks inter-subject and intra-subject analysis of the ERP.

It should be noted, however, that several runs with different stimulus parameters to gain more information on the brain responses of ANT DBS (not reported here) were conducted prior to the stimulation parameters used in the recording of the present study. These prior recording runs may have had a long-lasting influence on the brain networks, and hence on the ERP results presented in this paper.

## 5. Conclusions

We present a promising method for computing ERP related to ANT DBS and for ERP assessment. The applied stimulus parameters were 140 Hz monopolar and bilateral, which have not previously been studied on scalp EEG. The method is based on the optimization of stimulus on-off timing, signal preprocessing, epoch creation, EEG inverse imaging, and combined source and sensor level assessment. As a demonstration of the feasibility of the method, we analyzed scalp EEG during ANT DBS and uncovered a novel ERP-like response.

## CRediT author statement

**Herkko Mattila**: Conceptualization, Methodology, Formal analysis, Investigation, Writing – Original Draft. **Anna-Liisa Satomaa**: Formal analysis, Writing – Original Draft, Writing - Review & Editing. **Kai Lehtimäki**: Writing - Review & Editing. **Jari Hyttinen**: Writing – Review & Editing, Supervision. **Sari-Leena Himanen**: Writing – Original Draft, Writing - Review & Editing, Supervision. **Jukka Peltola**: Conceptualization, Writing – Review & Editing, Supervision, Project administration, Funding acquisition.

## Abbreviations

ANT: anterior nucleus of thalamus
DBS: deep brain stimulation
EEG: electroencephalography
DMN: default mode network
GABA: gamma-aminobutyric acid
MRI: magnetic resonance imaging
fMRI: functional magnetic resonance imaging
SEEG: stereo electroencephalogram
LFP: local field potential
ERP: event related potential
FIAS: focal impaired awareness seizures
ASM: anti-seizure medication
CT: computed tomography
EOG: electrooculogram
ECG: electrocardiogram
DC: direct current
IPG: implantable pulse generator
EMG: electromyogram
nTMS: navigated transcranial magnetic stimulation
FIR: finite impulse response
ICA: independent component analysis
BEM: boundary element method
dSPM: dynamic statistical parametric mapping
RTF: random field theory
SANTE: stimulation of anterior nucleus of thalamus for epilepsy
MR: magnetic resonance

## Data Availability

Research data is conditionally available from the authors with a formal data sharing agreement.

## Acknowledgements

This work was supported by the Research Council of Finland (Flagship of Advanced Mathematics for Sensing Imaging and Modelling grant 359185).

Tommi Nora and Mirja Tenhunen provided technical assistance during the EEG acquisitions. Antti Saastamoinen and Ella-Mari Salonen provided comments related to signal processing. Soila Järvenpää helped in the creation of Figure 1.

We thank François Tadel and the Brainstorm Forum for discussions about methodology and help with the Brainstorm software.

