## Supplement for "A method for analyzing the ERP associated with high frequency ANT DBS offset"

#### 1. Materials and methods

##### 1.1. DBS implantation

For the deep brain stimulation (DBS) implantation preplan, a 3-Tesla 0.9 x 0.9 x 0.9 mm magnetization prepared rapid acquisition gradient (MPRAGE) magnetic resonance image (MRI) (MAGNETOM Trio, A Tim System, Siemens Healthineers, Munich, Germany) of the head was acquired. In the MR images, the target anterior nucleus of thalamus (ANT) and the arteries to be avoided were defined. During implantation, the patient´s head was fixed to a Leksell stereotactic frame (Elekta, Stockholm, Sweden) and axial computed tomography (CT) (Revolution GSI, GE Medical Systems, Boston, MA, USA) imaging with slice thickness of 0.63 mm was performed. CT-images and preplan MRI were co-registered with Stealth Planning Station software (Medtronic Minneapolis, Minnesota, USA) to obtain a final plan with stereotactic coordinates of the target in the Leksell stereotactic frame system. A quadripolar DBS electrode (Model 3389, Medtronic Minneapolis, Minnesota, USA, Figure S.1.) was implanted bilaterally.

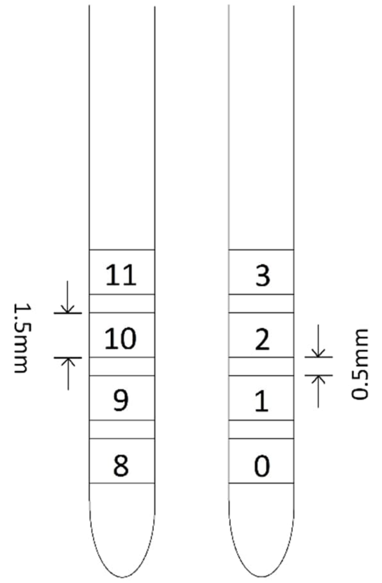


Figure S.1. Medtronic 3389 electrode. Electrode contacts 0-3 are on the left side and contacts 8-11 are on the right side. The length of each electrode contact is 1.5 mm and the spacing between each electrode is 0.5 mm.

The electrode contacts were numbered in inferior-superior direction 0, 1, 2, 3 on the left side and 8, 9, 10, 11 on the right side (Figure S.1). Contacts 2, 3 and 10, 11 were implanted inside the ANT,

(Figure 1). In the same operation, an implantable pulse generator (IPG) (Activa PC, Medtronic, Minneapolis, Minnesota, USA) was implanted subcutaneously below the subject´s clavicle.

### 1.2. Signal processing

#### 1.2.1. Event detection

The automatic event detection was done using an amplitude threshold detection for channel TP9. The parameters used in the Brainstorm software were a frequency range of 120 Hz to 160 Hz, and the threshold was set at 4 times the standard deviation. The stimulation onset events were detected in the same way but a frequency range of 10 Hz to 40 Hz was used, and the threshold was set to 4 times the standard deviation. To place the stimulation off and stimulation on event markers exactly in the middle of the first and the last stimulus pulse, respectively, a small time delay was manually applied to the events.

#### 1.2.2. ICA component removal

In the baseline session, two ICA components were removed to correct eye movement artifacts from the frontal channels. Small amplitude and continuous EMG artifact was reduced by the removal of four ICA components affecting channels AF4, F6, Fp1, FT9, and FP2.
In the stimulation session, six ICA components were removed to reduce eye movement artifacts from the frontal channels. Continuous small amplitude EMG artifacts were reduced by removing a total of 13 ICA components. The components' removal affected the following channels: F8(1), Fp1(3), Fp2(4), AF7(3), AF8(3), T7(1), TP7(1), F6(1), F4(1), C5(1), F5(1), F7(1), FC3(1), FC5(2), FT7(1), AF4(1), and C6(1), where the name of the EEG channel is followed by the number of the ICA components affecting that specific channel.
Nine ICA components (Figure S.3) were used to clean the edge effects of the stimulation artifact filtering.

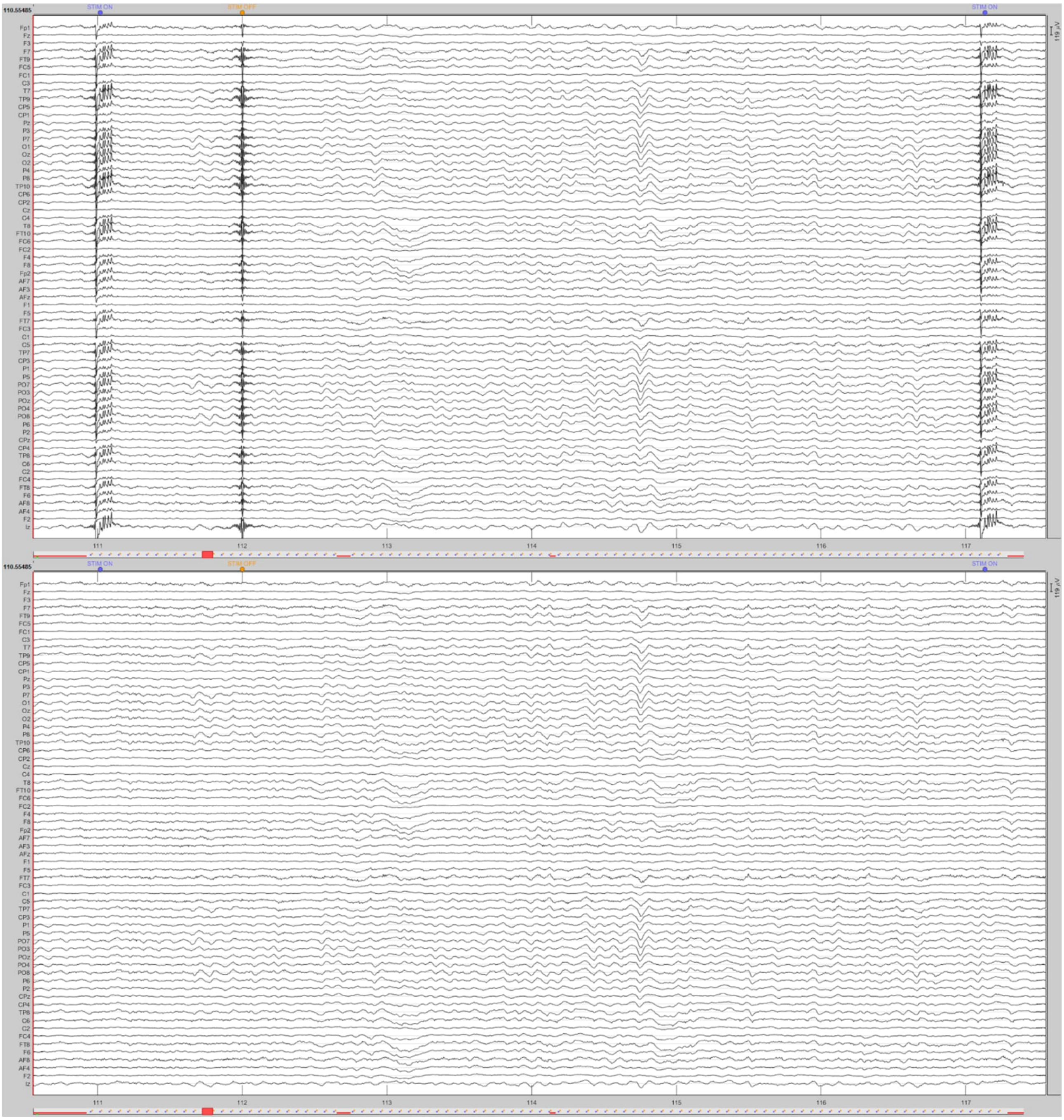


Figure S.2. Stimulation on and off segments in an EEG period of seven seconds. Upper image: the filtering artifacts at the beginning (onset) and the end (offset) of the 1 second stimulation train. Lower image: reduction of the filtering artifacts through the removal of the ICA component IC1.

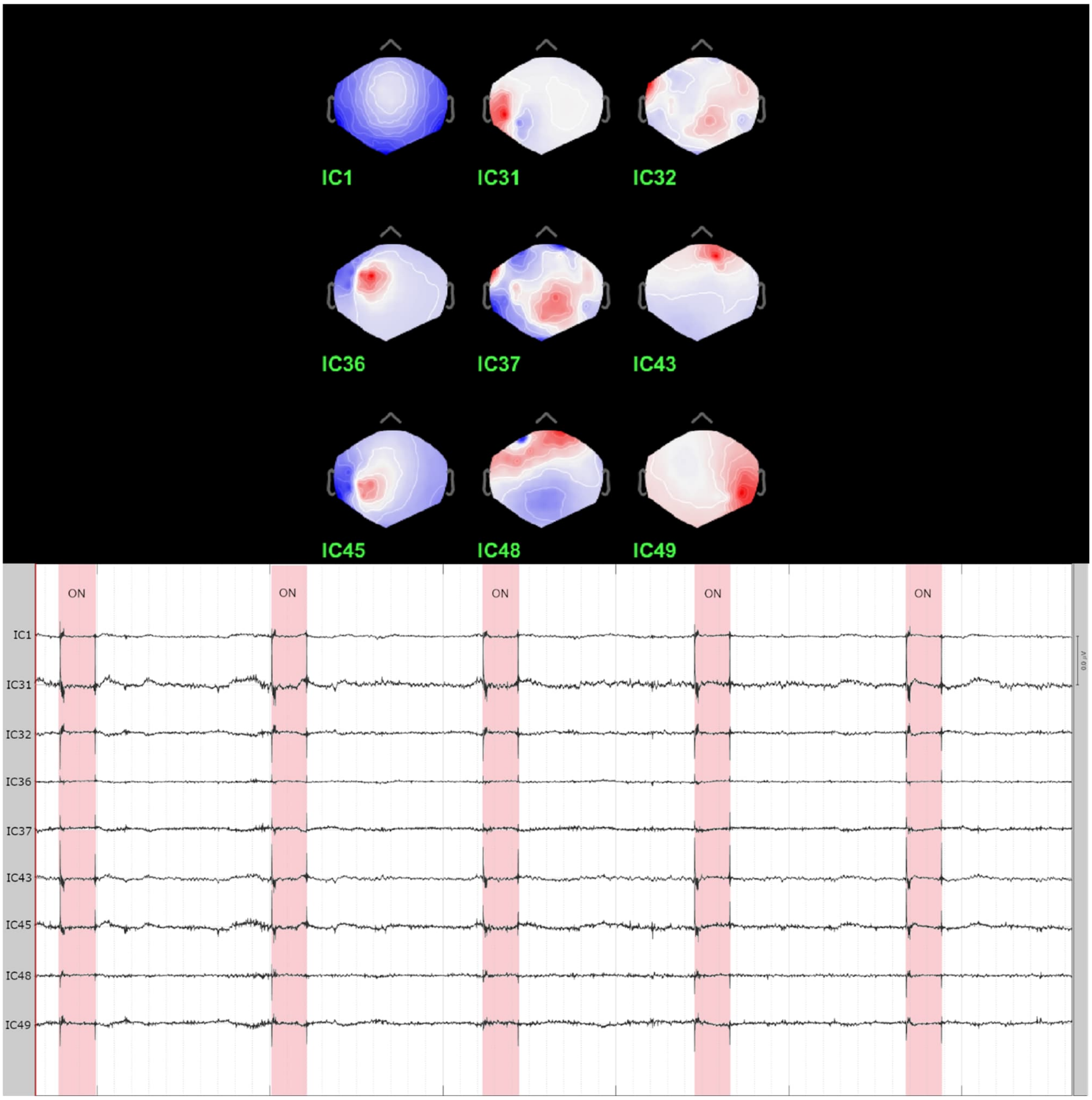


Figure S.3. Upper image: The topography of the ICA components used in the filter artifact correction. Lower image: example of the time series of the ICA components presented in the upper image.

#### 1.2.3. Investigation of genuineness of the ERP waveforms

It is possible that the ERP-like waveform seen in Figure 4 might have been caused by a band-pass filter transient of the stimulus artifact. In Figure S.4, the shape of the ERP-like waveform is observed in the unfiltered signal. It was also confirmed that the stimulus artifact did not saturate the amplifier, as no signal clipping was observed and the individual stimulus pulses were observed correctly. Therefore, the ERP-like waveform was not caused by a filter transient of the stimulation artifact or by the amplifier saturation post effects.

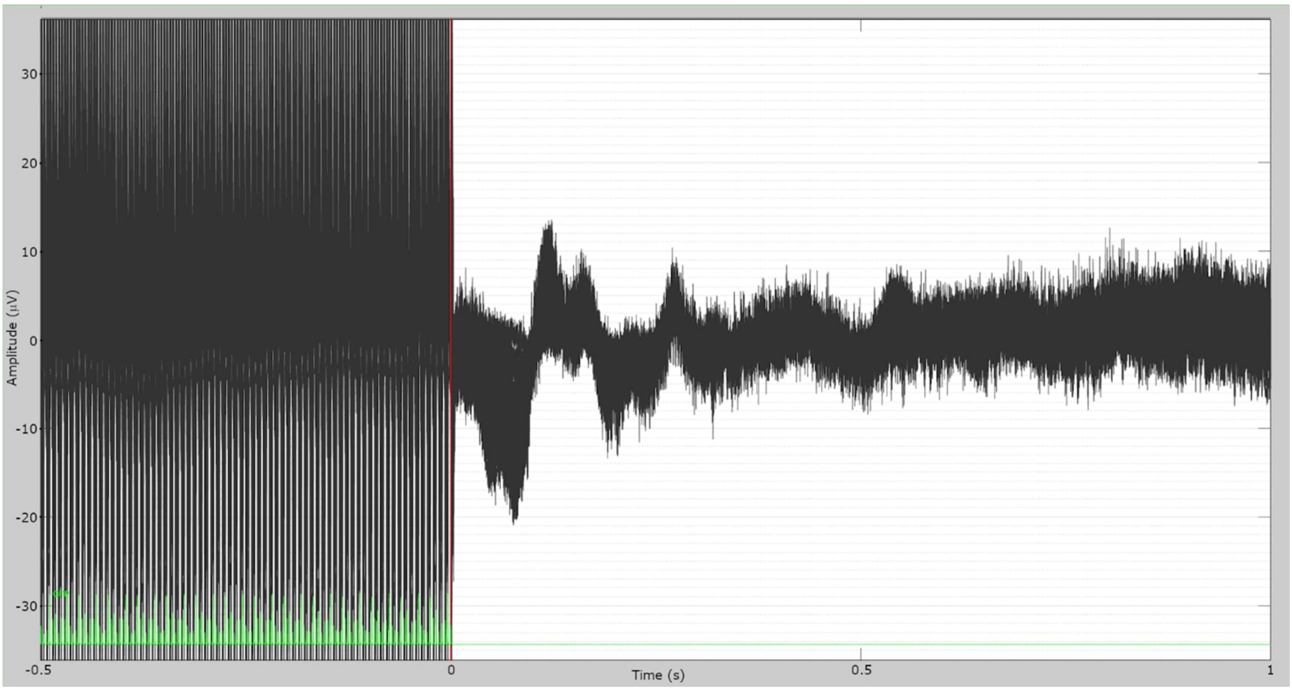


Figure S.4. Unfiltered raw data epochs averaged. The figure presents the same data as Figure 4 but without the 0.5 Hz to 100 Hz band-pass filtering. Butterfly plot of 64 channels. Total duration of the averaged epochs was 5500 ms, but only a time range from -500 ms to 1000 ms is presented. A high amplitude stimulus artifact can be seen in the time range of -500 ms to 0 ms. EEG presents an ERP-like waveform that takes place after the stimulus artifact even without filtering. The green curve in the bottom presents the GFP. The vertical scale of the figure is ±53.6 μV.

In addition, in Figure S.5 the data were epoched so that the whole stimulation artifact was left out of the epochs, and the data were 0.5 Hz to100 Hz band-pass filtered after epoching. The ERP-like waveform found was visually very similar to the waveform presented in Figure 4. Therefore, based on figures S.4 and S.5, it is very likely that the ERP-like waveform in Figure 4 represents true cortical activation.

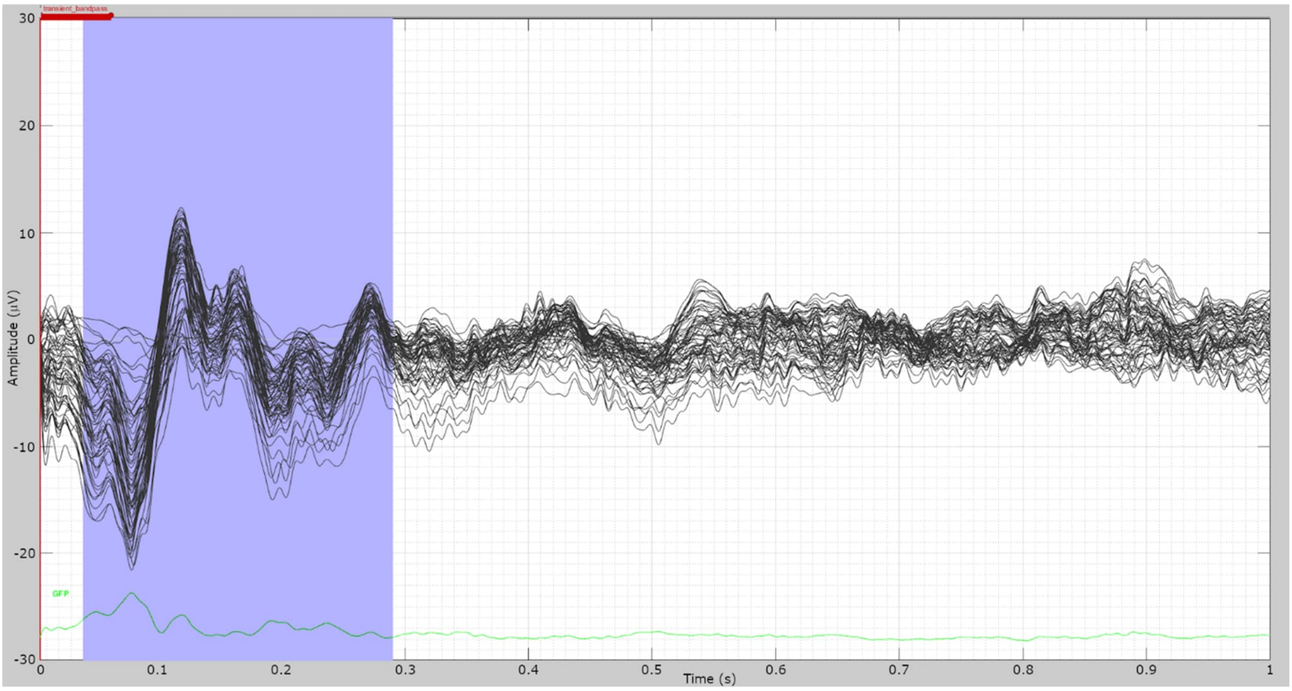


Figure S.5. Raw data epoched and averaged without the stimulus artifact. A 0.5 Hz to 100 Hz band-pass filter was applied after epoching and averaging. A butterfly plot of 64 channels in the time range of +5 ms to 1000 ms. The ERP-like waveform is visible in the time range of 40 ms to 290 ms (light blue area). The red bar in the upper horizontal axis denotes the band-pass filtering transient (0 s to 0.057 s) after the last stimulus pulse. This transient is observed because of the discontinuity of the signals in the first data points, and it might distort the data most strongly during the transient. The green curve at the bottom represents the global field power (GFP). The vertical scale of the figure is ±30 µV.

#### 1.2.4. Remainder sinc shaped artifact

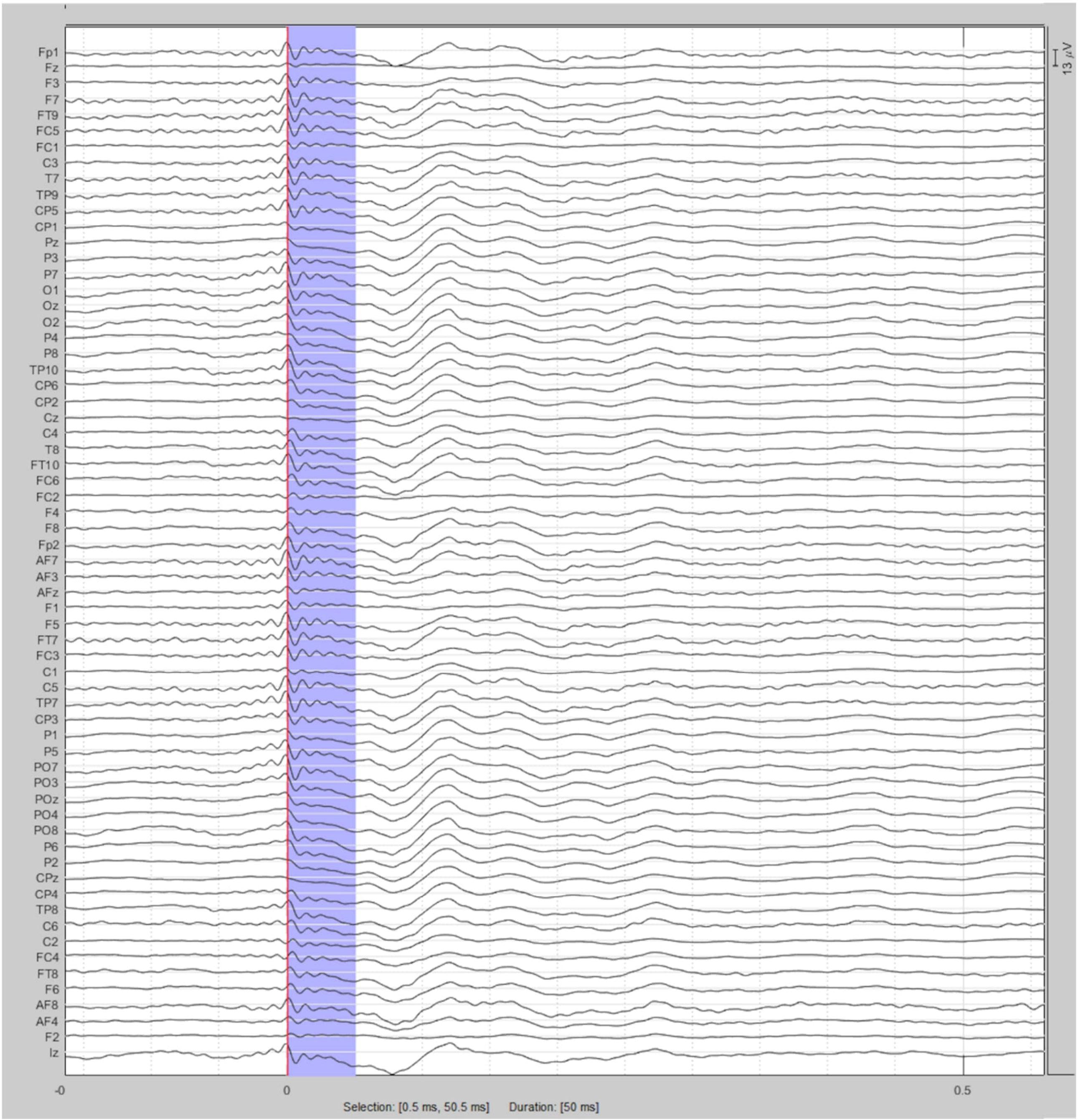


Figure S.6 The ERP response in the 64 EEG channels. The time range 0 ms to 50 ms is shaded light blue. There is a sinc shaped oscillation around the last stimulus peak caused by the EEG filtering. There was no obvious way to remove or clean this artifact. Therefore, the time range from 0 ms to 50 ms was not analyzed in the final analysis.

#### 1.3. Mixed source model

Similar to the surface model of the cortex, the automatic subcortical segmentation (Aseg) atlas created by FreeSurfer software was resampled to 15 000 vertices and the accumbens nucleus, amygdala, brainstem, caudate nucleus, hippocampus, pallidum, putamen and thalamus were bilaterally selected to the source model (Figure S.6).

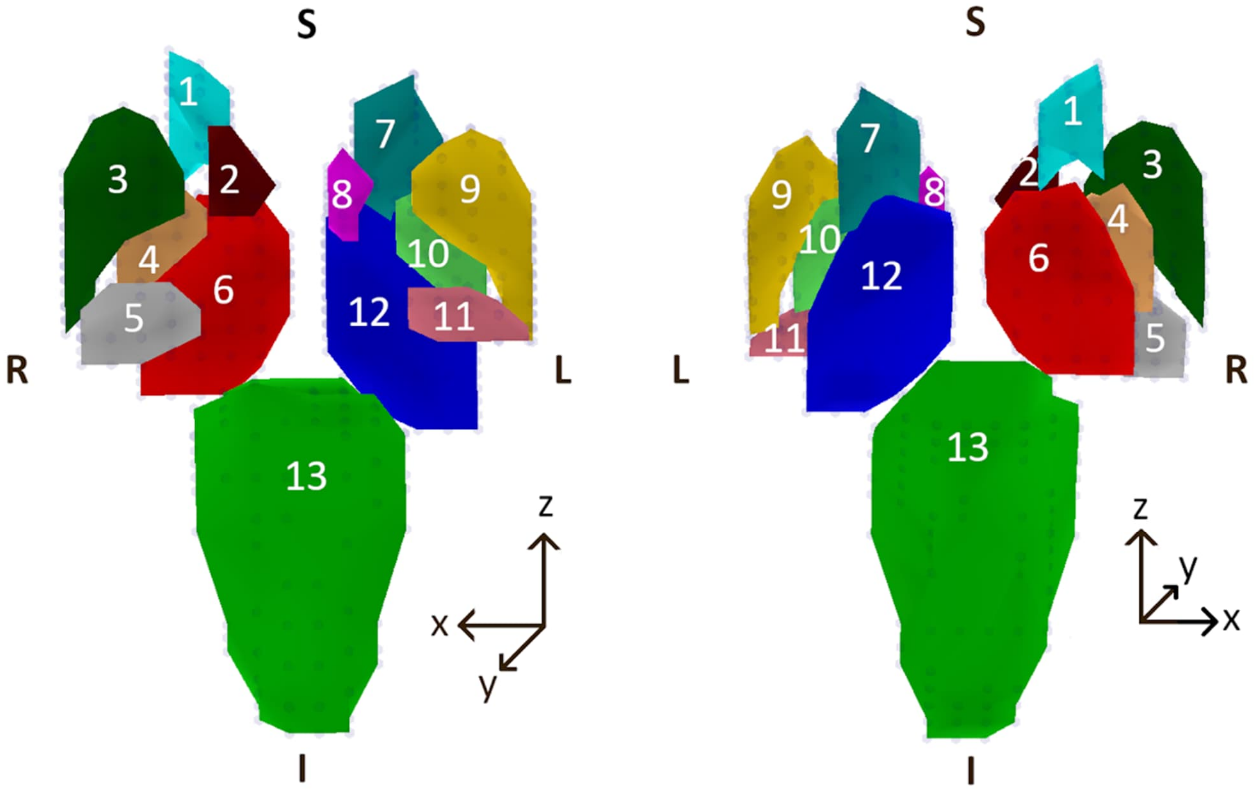


Figure S.7. Colored image of segmented subcortical structures and their orientation relative to the MRI. Anterior-posterior view (on the left) and posterior-anterior view (on the right). Right side: 1. caudate nucleus, 2. accumbens nucleus, 3. putamen, 4. pallidum, 5. amygdala and 6. thalamus. Left side numbering 7-12 is in the same anatomical order as the right side. 13. brainstem. Right (R), Left (L), Superior (S) and Inferior (I). The x-, y-, and z-coordinate axis of the MRI are presented in the lower right.

Cortical surface and subcortical structures were combined. In this new mixed source model, the cortex and the hippocampus were modeled as surfaces and rest of the subcortical structures were modeled as volumes (Attal et al., 2009; Attal & Schwartz, 2013). In the surface structures, one dipole was located to each vertex, pointing perpendicular to the surface (constrained model). All the volume structures, except for the pallidum, had unconstrained dipole orientation, meaning that three orthogonal (x-, y-, and z-directions) dipoles were located to each point of the volume grid. In the pallidum, dipoles in a volume grid were constrained to the y-axis direction (Figure 4) (Attal et al., 2009).

### 2. Statistically significant Deep brain activity

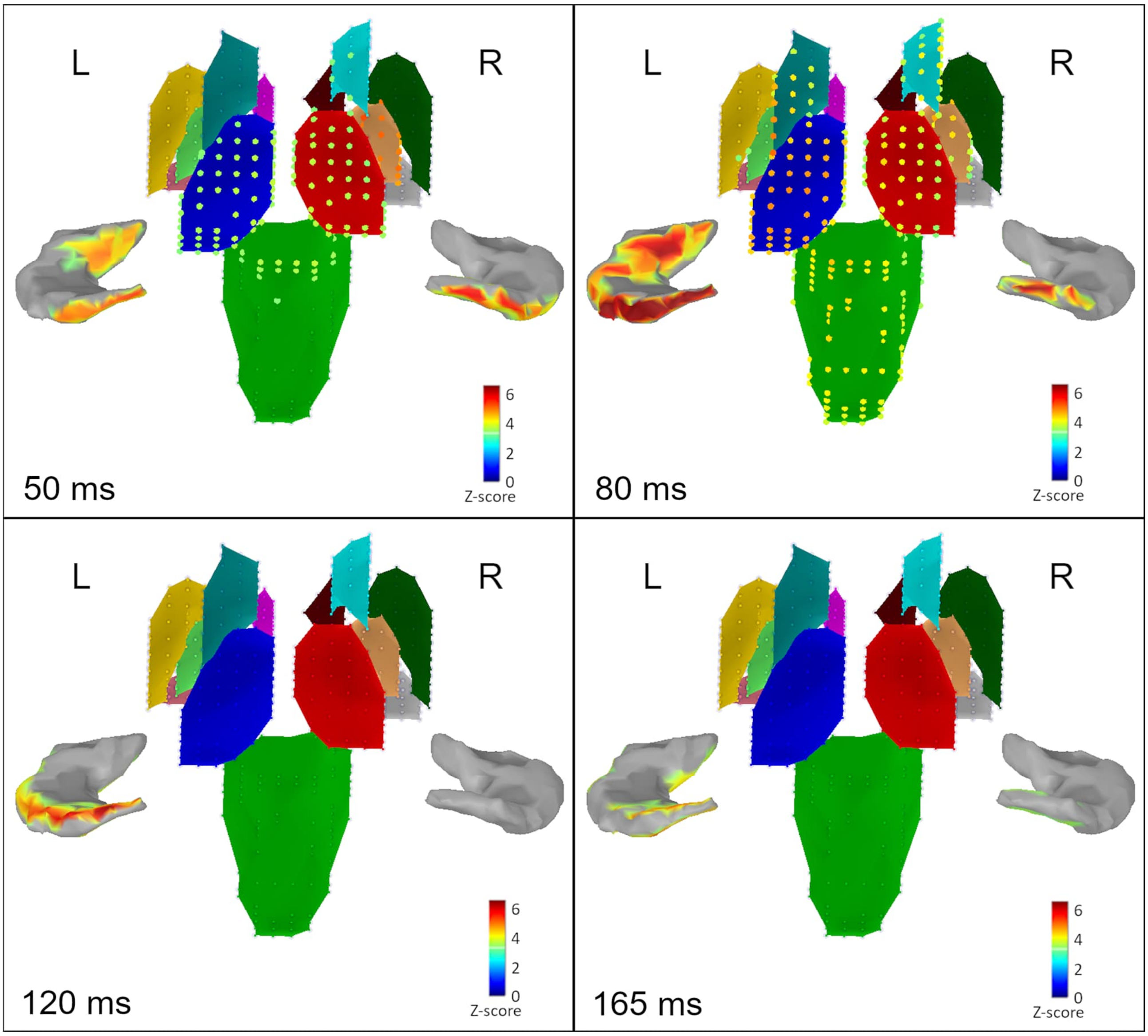


Figure S.8. Activity of the subcortical sources and hippocampus. The hippocampus presents the surface model activations, whereas the volume grid points present activations of the subcortical structures: the caudatus nucleus, accumbens nucleus, putamen, pallidum, amygdala, and thalamus. The structures and their colors are the same as those presented in Figure S.7. The direction of view is posterior to anterior. Upper left P50, Upper right P80, lower left N120, and lower right N165. Activity with $p<0.001$ is presented and activity was observed in the hippocampus, caudate nucleus, pallidum, thalamus, and brainstem.
